# Reach, adoption, and implementation strategies of a telehealth falls prevention program: perspectives from francophone communities across Canada

**DOI:** 10.1101/2023.09.27.23295569

**Authors:** Jennifer O’Neil, Nathalie Dionne, Sylvie Marchand, Dominique Cardinal, Grant Handrigan, Jacinthe Savard

## Abstract

**Introduction:** A fall may impact a person’s physical, emotional, and psychological well-being. Fall prevention programs are being implemented to reduce these negative outcomes. However, linguistic barriers in health services may reduce access to such prevention programs. A telehealth fall prevention program was designed to increase access to such programs in French for Francophone minority communities in Canada. This capacity building project aimed to support community partners to deliver this telehealth program and document strategies used to reach, adopt, and implement the program within various Francophone and Acadian Minority Communities.

**Methods:** A sequential explanatory mixed methodology was used to document reach, adoption and implementation strategies and describe lived experiences of program facilitators and organization representatives. Reach, adoption, and implementation were documented and analyzed descriptively while lived experiences were analyzed using content analysis following the Consortium Framework Implementation Research.

**Results:** Twelve organization representatives or program facilitators from eight organizations operating in four different provinces participated in the study. Three themes emerged from the qualitative data on reach and adoption: external context, internal context, and capacity building. Four themes were identified as barriers and facilitators to implementation: level of preparation and time management, interpersonal relations and telepresence, exercise facilitation and safety, and technological problem-solving.

**Conclusion:** Using tailored reach and adoption strategies such as prioritizing provinces with higher proportions of needs and training local community program facilitators may lead to successful implementation of a new telehealth fall prevention program. Results from this study could potentially inform other primary prevention programs or telehealth program implementation.

## Introduction

A fall can lead to a significant injury which may impact a person’s physical, emotional, and psychological well-being. Falls in older adults are the leading cause of injury or emergency visits in Canada. Current evidence around multifactorial fall prevention programs that focuses on people presenting in the emergency department due to a fall does not demonstrate a significant impact on fall incidence (Morello et al., 2019). Therefore, the implementation of programs for people at risk of falls before they need medical care should be further explored.

Linguistic barriers in health services may lead to delaying/reducing preventive consultations, poor health outcomes and poor health service quality (Bowen, 2001, 2015; de Moissac & Bowen, 2019). Health interventions in French for Francophones may improve confidence and the therapeutic relationship (Drolet et al., 2015), two critical aspects of intervention effectiveness. Therefore, implementing fall prevention programs tailored to older adults in their language of choice may be a solution to reduce falls among francophone older adults living in minority situations. Telehealth increases the potential to reach small or remote communities in which local French speaking healthcare professionals are not available.

With an overarching goal of improving access to health services in French, the telehealth fall prevention program *Marche vers le futur (MVF)* is an effective primary fall prevention program prioritizing francophone adults 55 years and older, who present with fall risk factors (Savard et al., 2018). This multifactorial program including pre-post assessments and a 10-week intervention was developed for Francophone and Acadian Minority Communities (FAMC).

Implementation literature reports several factors capable of positively or negatively influencing the implementation success of a new health program within a health optimization. These are the presence of local leaders, the social influence and local context (Goorts et al., 2021). However, there is limited research around implementation strategies in the context of health primary prevention in FAMC or regarding telehealth programs that incorporates multifactorial components (i.e., education, exercises, and discussions). This study will provide information on lived experiences and the strategies used by Francophone organization representatives and program facilitators to reach communities, improve adoption, leading to successful implementation of the MVF program across FAMC. This study aims to document 1) reach - defined as the potential individuals and eligible communities, 2) adoption - focusing on internal and external contexts and reasons for adoption, and 3) implementation - perspectives from program facilitators.

## Methodology

A sequential explanatory mixed method was used to document the strategies used by organization representatives and program facilitators and lived experiences of implementing the telehealth MVF program. The conceptual frameworks RE-AIM (Kwan et al., 2019) and its extension PRISM (McCreight et al., 2019; Trinkley et al., 2020) were used to guide data collection. The Consortium Framework Implementation Research (CFIR)(Damschroder et al., 2009) was used to inform data analysis. External validity was established by using the RE-AIM checklist to report information on reach and adoption. The Good Reporting of A Mixed-Method Study (GRAMMS) checklist as well as the six recommendations to improve adequate reporting of the methodology (Lengnick-Hall et al., 2022; O’Cathain A et al., 2008) was also used to enhance rigor in reporting on implementation.

### Participants and sampling

Purposeful sampling was used to recruit organization representatives and program facilitators in FAMC interested in providing health prevention services to Francophone older adults in their communities. Each organization representative or program facilitator who adopted the program was invited to participate in the study and signed a consent form prior to completing any questionnaires.

### Intervention

The MVF falls prevention program is delivered remotely, tailored, and effective in reducing fall risk factors in francophone adults aged 55 and over. Each 90-minute session offered by a trained program facilitator once a week, for 10 weeks includes an educational component and discussion around fall risk factors followed by an exercise program targeting balance, strength, and flexibility. Details of the program can be found in Savard et al. (Savard et al., 2018) and in Appendix1. In this study funding was available for the community agency who wanted to implement the project. Two versions of the program were offered: MVF community where participants met in a local center to complete the program and MVF home where participants connected virtually from their residence.

### Demographic and quantitative data

Demographic data including gender identity, language preferences, age and professional experience were used to contextualize the group of animators and organization representatives. Reach, adoption, and implementation data were documented before, during and after program implementation. Reach was measured as the number of the Francophone community contacted by the team, the number of provinces involved, and the number of program facilitators recruited.

Adoption was documented by the number and percentage of communities implementing the program, and the setting of each community. Implementation was documented descriptively by documenting the number of participants who completed the assessments, who participated in the program, as well as attrition. Representatives’ and program facilitators’ perspectives on the implementation were also documented.

### Qualitative data

Data reporting on lived experiences related to the reach of organization representatives and program facilitators, and adoption of the program were documented using individual interviews. Implementation of the program was documented using field notes. An MVF trainer (ND) attended the first, second and sixth session in person or virtually to document program implementation through field notes for each community.

### Data analysis

A mixed method of analysis was used to triangulate the data. Demographic data were analyzed descriptively (i.e., percentage, mean, standard deviation). Quantitative data was categorized between reach and adoption and analyzed descriptively (i.e., percentage, mean). Pre-program implementation perspectives were compared with post-program results using the CFIR framework and using implementation outcome to document if implementation expectations were under- or overestimated.

Qualitative data was analyzed using content analysis following the CFIR framework. Field notes were deductively organized by types of strategies (e.g., reach, adoption, or implementation). Furthermore, the field notes documenting the progress of the program facilitator over the 10 weeks program implementation were deductively categorized under barriers or facilitators. Data from the interviews was transcribed and inductively coded into new themes by two physiotherapist-researchers involved in the implementation of the MVF. The lived experiences of the program facilitators and organization leaders was reviewed by the research team including occupational therapists, physiotherapists and MVF program developers. The quantitative and qualitative data was then integrated in the discussion to allow contextualization of the information.

## Quantitative Results

### Demography

Eight organizations from four provinces, three organization representatives and ten program facilitators consented to participate in this study. One organization representative dropped out due to family reasons. Of the twelve organization representatives or program facilitators who completed the study, most were francophone females born in Canada and identifying as cisgender ranging between 20 and 59 years old without previous experience implementing a telehealth program. Five health professions were represented and included physiotherapy (25%), occupational therapy (25%), kinesiology (17%), physical activity or specialized education (17%), social work (8%), and retired healthcare workers (8%) (table 1). Together they implemented 16 MVF programs, representing one to three implementations per community (Table 1).

**Table 1.**
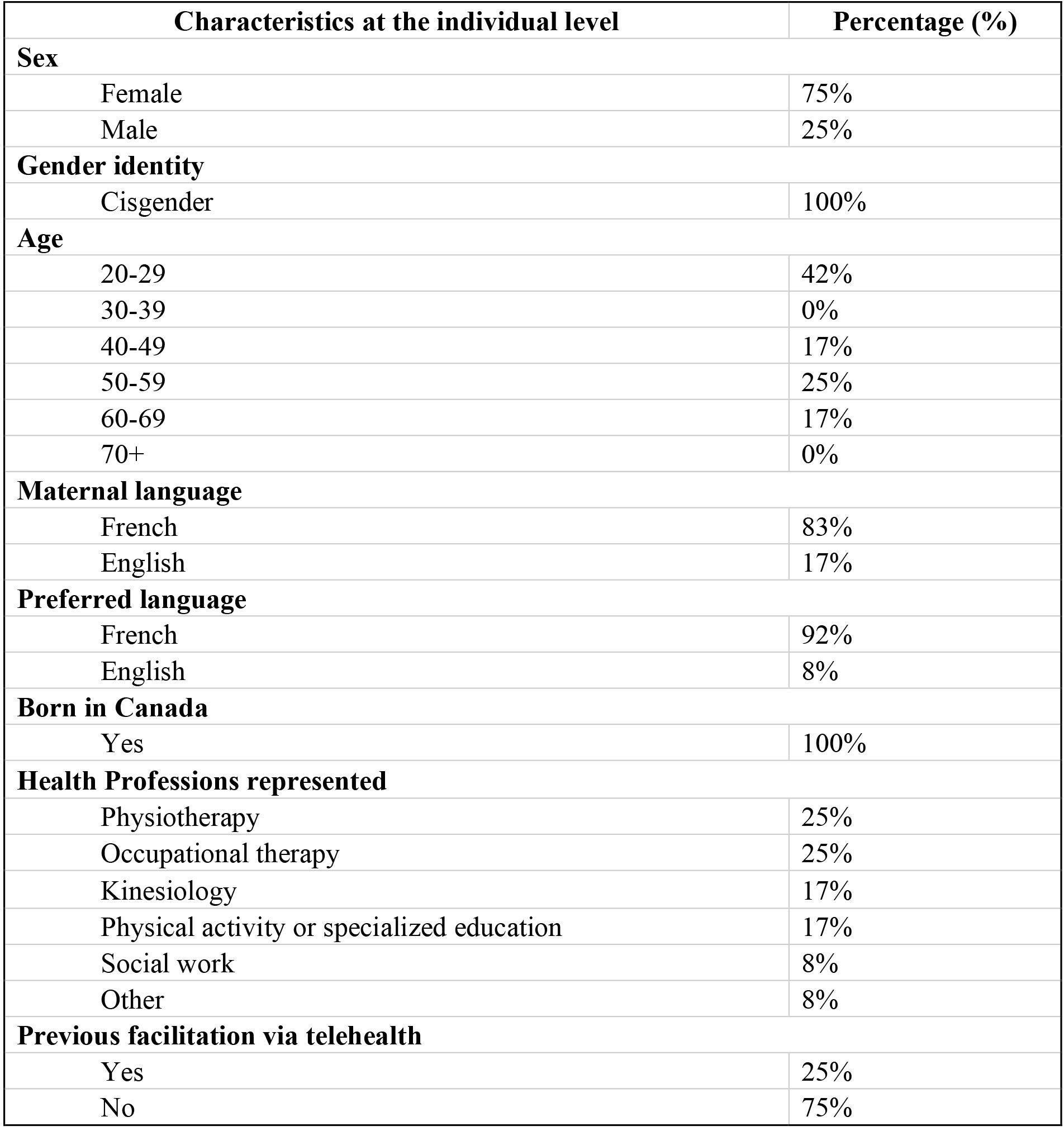
Organisation representatives and program facilitators’ characteristics.

### Setting characteristics: Reach

Thirty-four Francophone communities were contacted between June 2021 and September 2022, using three communication modalities such as email, phone calls and online meetings, for an average of 5 to 10 hours per week allocated to relationship building. Reach and recruitment efforts were also supported by a promotional video and an infographic presenting the program. Each community was contacted by the same person multiple times by email, followed by a phone call or online meetings. Phone calls or online meetings ranged between 20-60 minutes per community. The median age of the francophone population in the contacted provinces who responded (NB, AB, ON, NS, PEI, Nunavut, YK, TNO) ranged between 37 and 55.2 years old, and the percentage of individuals declaring French as their first official language spoken range between 0.5% and 31.8% (table 3).

### Setting characteristics: adoption

Out of 34 communities contacted over one year, 8 organizations from 4 different provinces adopted the telehealth program (24%). Hospitals, community health centres and community programs respectively represented 13%, 25%, and 63% of the settings in which the program was adopted (Table 2). Overall, 33 program facilitators were trained over 10 two-day courses between December 2021 and February 2023. Of the provinces which implemented the program, one facilitator was trained in Alberta, six in New Brunswick, four in Prince-Edward Island, and eight in Ontario.

**Table 2.** Characteristics of contacted communities (i.e., percentage (%) of francophones living in the community, median age of the francophone adults, percentage of older francophone adults, and rural-urban proportions) Characteristics of community who adopted the program MVF are in bold. These statistics were taken from the Canadian Heritage Official-Language Minority Communities Dashboard (2021).

| Francophone communities in each province-territories | Francophones living in province (%) | Median age of francophone adults (years) | Proportion of older francophone adults (60 years and over) (%) | Proportion of francophones living in Rural/Urban Areas (%) |
| --- | --- | --- | --- | --- |
| British Columbia | 1.4 | 51.1 | 30,8 | 13.2/86.8 |
| **Alberta** | **2** | **43.9** | **21,7** | **16.1/83.9** |
| Manitoba | 3.2 | 51.4 | 32,8 | 27.8/72.2 |
| Saskatchewan | 1.3 | 55.2 | 39 | 35.1/64.9 |
| **Ontario** | **4.1** | **47.9** | **27,1** | **17.8/82.2** |
| **New Brunswick** | **31.8** | **48.8** | **29,9** | **51/49** |
| Nova Scotia | 3.2 | 53.8 | 36,9 | 58.5/41.5 |
| **Prince Edward Island** | **3.3** | **52.4** | **34,1** | **55.3/44.7** |
| Yukon | 4.6 | 41.1 | 16,8 | 17.6/82.4 |
| Northwest Territories | 3 | 37.5 | 9,7 | 100/0 |
| Newfoundland and Labrador | 0.5 | 45.4 | 25,9 | 45.8/54.2 |
| Nunavut | 1.8 | 37 | 10,5 | - |
| Total Francophones outside Quebec | 3.8 | 48.4 | 28.1 | 27.7/72.9 |

### Setting characteristics: implementation

A total of 146 older adults were assessed and 129 completed the program (88%). On average, 2.4% attrition was documented (n=7) following the initial assessment.

### Pre implementation perspectives from program facilitators compared to the reality

Program facilitators who responded to the preprogram questionnaire (n=7) slightly underestimated the potential recruitment of older adults, estimating 14 participants (± 3.14) compared to the reality which averaged 18 participants per program. As for the percentage of older adults accepted into the program, facilitators underestimated the acceptance rate in the preprogram questionnaire, estimating 82.9% compared to the reality with 93% of older adults accepted. Regarding the type of telehealth service provided, program facilitators accurately anticipated the location of the MVF telehealth program, estimating 80.7% of the program delivery in centers and 22.1% at home compared to an actual 80,8% and 19,2% respectively.

## Qualitative perspectives from community representatives and program facilitators

### Reach and Adoption

Reach and adoption data were categorized into external context, internal context, and capacity building. Three sub-themes emerged in the external context category: pandemic-related barriers, infrastructure, and regional priorities. Community representatives and program facilitators reported that public health restrictions caused delays in adopting the new program.

This was mostly related to limited human resources including limited Francophone healthcare professionals capable and willing to facilitate the MVF program. Program facilitators and representatives shared that the MVF program allowed them to reduce isolation, offer a service in French, increase physical activity in older adults, and fulfill a gap around fall prevention Francophone programming in their region. Technology literacy was a limiting factor in the program’s adoption for some communities, and limited financial support was also reported as a barrier for long-term implementation limiting initial adoption.

Regarding internal context, compatibility of the telehealth falls prevention MVF program with the organization’s strategic priorities was described as important for most communities. However, organizational, and structural barriers including multiple decisional levels, complex financial procedures or limited partnerships in the community were reported as challenges. Even though financial support from Health Canada/Société Santé en français (SSF) allowed the project team to support the salary of all program facilitators and acquisition of equipment, challenges with creating effective financial procedures were raised. Many community organizations did not have the financial capacity to purchase equipment and afford salaries prior to being reimbursed, which caused important barriers.

Ongoing support and communication through the pre-implementation steps from the MVF research team was expressed as being critical for capacity building. Each community interacted with the MVF team monthly for 6 to 12 months before implementing the program. This may have facilitated the program adoption.

### Implementation

Four themes were identified by the program facilitators as barriers and facilitators to implementation: level of preparation and time management, interpersonal relations and telepresence, exercise facilitation and safety, and technological problem-solving.

Preparation before each class included looking back on previous weeks and introducing the week’s topic. Insufficient planning by several of the facilitators was observed by the MVF trainer in the first sessions, but only from one facilitator by week six demonstrating learning in this area. For time management, program facilitators initially underestimated the length of the exercise program. However, all facilitators were able to improve and deliver the entire content of each class within the planned time by week six.

From an interpersonal relationship perspective, several facilitators showed signs of nervousness, and some were lacking in energetic disposition during the first few classes. Remote interaction with participants sometimes lacked fluidity. Some facilitators initially forgot to engage with participants virtually or were too far from the screen. However, with feedback by week six, all facilitators were interacting with more ease, calling participants by their first names, feeling more comfortable with group discussions, and encouraging participants to talk amongst themselves. To stimulate motivation in participants, some program facilitators used encouragement techniques such as verbal cueing, counting repetitions out loud and words of encouragement from the first session. Telepresence concepts such as optimal positioning in front of the screen and wearing contrasting clothing to maximize visibility were well demonstrated from the outset.

When demonstrating exercises, a lack of clarity in the instructions occasionally caused some confusion for participants. Improvement in this area was observed at all sites during subsequent classes. Safety (i.e., wearing appropriate footwear for the exercises, correcting participants as needed, giving advice on how to modify the exercises when some had difficulty) was well assured by program facilitators throughout the program. Infrastructure selection for community programs, including lighting, room size and layout was adequate in most cases. Only one facilitator needed safety reminders during week six.

Occasionally unstable internet connection and lack of technological preparation (i.e., uncharged computers, missing charging cables, access to password protected on-site computers) posed some challenges. Despite the occasional technological limitations, all program facilitators showed good technological problem-solving skills to ensure that all sessions could be completed smoothly. For example, several facilitators connected to the virtual session on a second device such as a laptop or tablet, placed close to them allowing them to demonstrate exercises while observing participants adequately. Program facilitators also reported that the additional training and feedback received during the implementation were valuable to increase their comfort level in delivering the program.

## Discussion

This study documented strategies to reach, adopt and implement a new telehealth program for primary fall prevention according to the lived experiences of program facilitators within various communities.

Since building community partnership is essential for the successful adoption of new health interventions (Mott et al., 2014), dedicating time and human resources to build these partnerships was vital to the implementation of the MVF program. Our results demonstrated that an entire year was dedicated to reaching potential Francophone communities and building partnerships. Community reach was completed by the same researcher, a physiotherapist with a PhD in rehabilitation, and expertise in telerehabilitation and fall prevention as well as lived experience animating and implementing the telehealth fall prevention program MVF.

The belief and knowledge that fall prevention should be prioritized within each organization may have facilitated adoption. Supporting the literature (Sévigny et al., 2015), results from our study shows that meaningful engagement of organizations with a mission to improve access to health services for Francophone adults, and ongoing communication with program facilitators may have supported adoption and facilitated implementation. Organizational readiness has been documented as a key factor in the implementation of new evidence-based programs (Scaccia et al., 2015). Concordance between the mission and mandate of each organization and the objectives of the MVF telehealth fall prevention program allowed for a seamless adoption of the program since community organization were ready.

In addition to the mission of an organization, targeting the right context and demographic is critical to the adoption of a new program. An awareness of the local region is necessary for implantation to be successful. Results from our study demonstrated that program facilitators underestimated the interest for the program, and acceptance rate of older adults into the program. Interestingly, two out of four provinces who adopted the program were amongst those with the highest rural to urban living proportions. This would suggest that having access to various versions of a telehealth program may facilitate recruitment for people living in rural areas.

However, further efforts should be made to recruit participants from provinces where the population aged 60 and over represents more than 30% of the francophone population (British Columbia, Manitoba, Nova Scotia, and Saskatchewan).

The use of telehealth may facilitate reaching remote communities in which local French-speaking healthcare professionals are not available. Perspectives around technology literacy were reported by facilitators which concord with the current literature around the importance of telepresence and digital literacy (Triana et al., 2020). Experience using technology to provide health services is crucial to the success of a program. Our results demonstrated that facilitators’ skills and comfort with the technology improved over time and that the presence of the MVF trainer during the implementation was a key factor in this improvement. Certain difficulties were mitigated by good preparation of the facilitators and continued education throughout the implementation which included how to demonstrate exercises while observing and providing corrections to each participant; and improving their familiarity with the content. Many facilitators seemed to have underestimated the preparation required to teach the exercises with a comfortable level of confidence.

### Limitations

Our results reflect only the reach, adoption, and implementation of MVF by eight FAMC from four provinces. Future studies should focus on reaching communities from provinces and territories with a smaller pool of Francophones to improve access. The financial support from SSF/Health Canada for this project allowed communities to acquire equipment, training, and support for the program implementation; therefore, transferability to other contexts when such financial support is not available may be limited. The authors are aware that the interpretation of the data may be biased as they were involved in supporting various program implementation, but their positionality statement may offer readers a better understanding of how the data was interpreted, therefore improving the trustworthiness of their interpretation.

### Key implications for public health interventions, practice, or policy

Time and facilitator engagement are factors well reported in the literature as essential components to successful implementation (Boothroyd et al., 2017). Results from our study support this and contribute by reporting on the importance of capacity building led by a professional with lived experience. Assessment of an organization’s readiness should include fit between the organization’s mission and prevention program and adequate timing of the implementation. Workshops for all stakeholders involved in a new telehealth program should include how to use of specific platforms, how to interact with your audience using technology, how to problem solve different scenarios and a simulation component before engaging with the participants.

## Conclusion

Networking, credibility, effectiveness, and access to financial resources are necessary strategies to facilitate the reach, adoption, and implementation of a novel telehealth primary fall prevention program. Similarly, establishing the availability of Francophone human resources by training community health professionals, securing proper infrastructure and financial support before implementation is critical. Finally, using tailored reach strategies such as focusing on provinces with higher proportions of older Francophone adults or rural regions where Francophone health services are limited and training local community program facilitator may lead to successful adoption of a new telehealth fall prevention program. To ensure sustainability of the program, a community of practice will be created to provide a space for program facilitators to exchange their experiences and ask questions. The shared experiences and documented strategies used by healthcare providers in Francophone and Acadian Minority Communities may guide other community healthcare workers, clinical managers, and healthcare organizations in their implementation of primary prevention programs delivered using telehealth.

## Data Availability

All data produced in the present study are available upon reasonable request to the authors

## Acknowledgements

This study was approved by the UOttawa research (H-11-21-7449), the Bruyère Research Institute (M16-22-016), and the uMoncton board of ethics (2122-090).

